# PlanetDiet: Sustainable diets and cardiometabolic health, study protocol for a six-month parallel-group randomized controlled trial in middle-aged adults at cardiovascular risk

**DOI:** 10.64898/2026.11.06.26367228

**Authors:** Marta Trius-Soler, Teresa Costa-Seco, Adam Rewers, Catalina Cuparencu, Anne-Marie Raabyemagle, Inge Tetens, Marta Guasch-Ferré

## Abstract

**Background:** Cardiovascular disease (CVD) remains the leading cause of death worldwide. Diet plays a key role in CVD prevention while linking human health with environmental sustainability. The EAT-Lancet Commission proposed the Planetary Health Diet (PHD), a predominantly plant-based dietary pattern emphasizing high-quality foods and reduced meat consumption. Despite growing interest in the PHD, no randomized controlled trial (RCT) has evaluated its effects using the PHD index on established and novel cardiometabolic biomarkers.

**Methods:** The PlanetDiet study is a six-month, three-arm parallel RCT investigating the effects of sustainable diets on traditional and novel cardiometabolic risk factors in 180 Danish adults aged 45–70 years at high cardiovascular risk. Participants are randomized to a PHD, an ovo-lacto-vegetarian diet, or their habitual diet (control). The primary outcome is changes in a composite cardiometabolic health score, with secondary outcomes including changes in metabolomic and proteomic profiles. Clinical assessments include the analysis of urine, blood, and faecal samples; continuous glucose monitoring; anthropometry and body composition measurements; dietary records; and self-reported questionnaires assessing physical activity, quality of life and dietary acceptance. To support adherence, participants receive four individualized dietary counselling sessions, two group sessions, seasonal food boxes tailored to their assigned diet, and access to a personalized digital application with recipes, individualized timelines, and visit scheduling.

**Conclusions:** PlanetDiet will provide evidence on the effects of sustainable dietary patterns on cardiometabolic health. It will also investigate molecular pathways linking diet to cardiometabolic disease. Ultimately, the study may inform the development of effective, personalized dietary interventions that promote both human and planetary health.

Trial registration: Sustainable Diets and Cardiometabolic Health (PlanetDiet): NCT07189676 [<u>ClinicalTrials.gov</u>] [registered before inclusion start; 2025-09-15]; Unique identifier: H-25030722.

**Graphical abstract:** 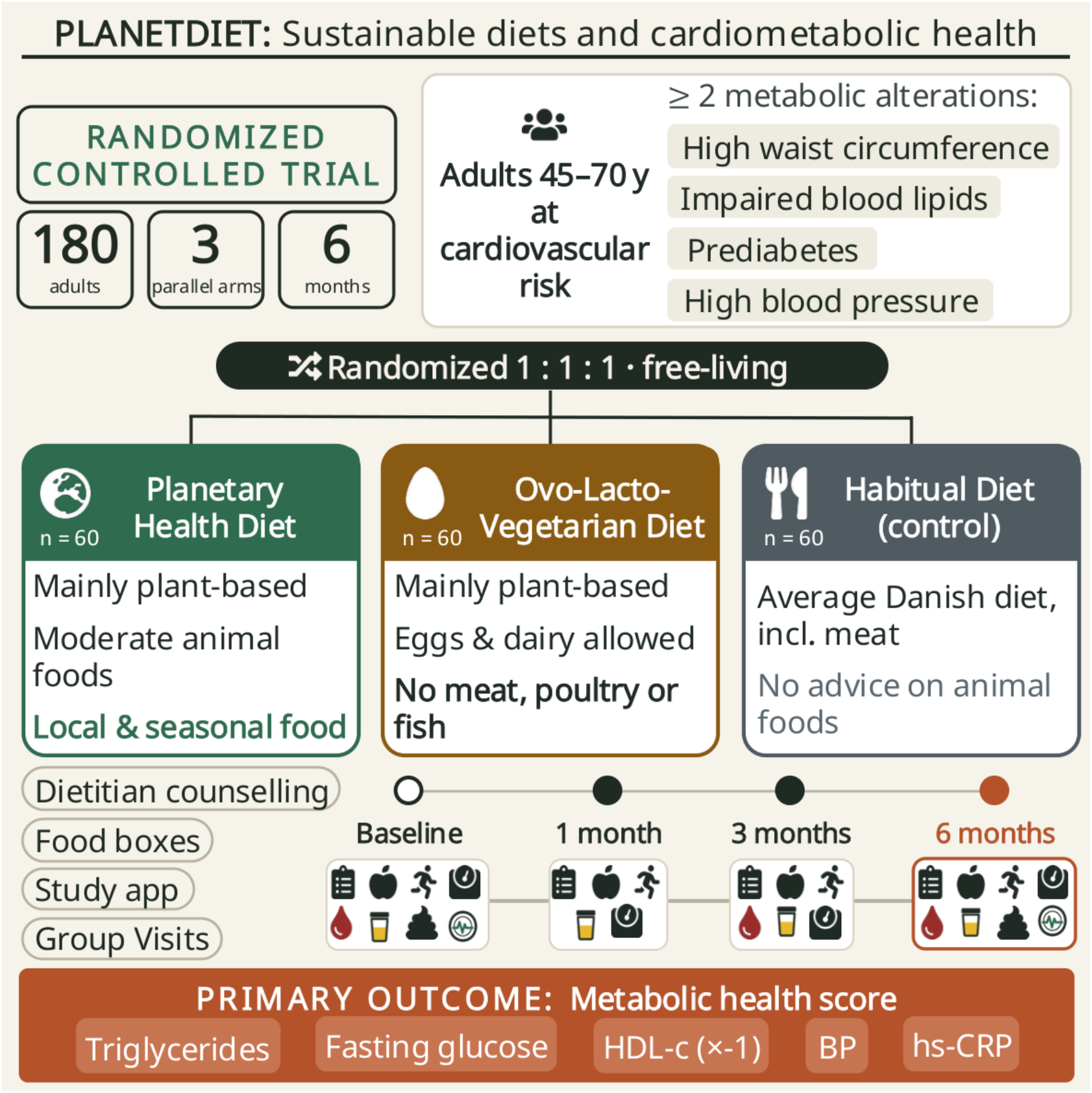

**Clinical perspective:** Randomized controlled trials are needed to test the effects the Planetary Health Diet EAT-Lancet framework on cardiometabolic health markers.

The PlanetDiet trial will provide insights into the feasibility of implementing sustainable diets and achieving participant adherence in a middle-aged population at cardiovascular risk.

Omics approaches could identify intervention-responsive molecular features and generate hypotheses about pathways linking plant-based dietary change to cardiometabolic health.

## Introduction

Diet is a cornerstone of cardiovascular disease (CVD) prevention ^1^. Epidemiological studies and clinical trials have shown that diets high in red and processed meats are associated with increased risk of type 2 diabetes and CVD^2–4^; whereas high-quality plant-based diets are associated with lower risk of CVD and mortality ^5–7^. Beyond their implications for human health, dietary choices and food systems have substantial environmental consequences ^8^. Animal agriculture, particularly beef production, contributes to greenhouse gas emissions, land-use change, biodiversity loss, and nitrogen pollution ^9,10^. These interconnected challenges have prompted official dietary guidelines worldwide, including those in Denmark, to integrate environmental sustainability with recommendations for health promotion ^11^. Evaluating dietary patterns that address both human and planetary health is therefore increasingly relevant to CVD prevention.

In 2019, the EAT Lancet Commission called for a global dietary transformation by 2050, advocating a shift toward dietary patterns that align food systems with environmental sustainability and human health ^8,10^, while also supporting the United Nations Sustainable Development Goal. The Commission proposed the Planetary Health Diet (PHD), a predominantly plant based dietary pattern that emphasizes whole grains, vegetables, fruits, nuts, and legumes, allows limited amounts of dairy, eggs, seafood and poultry, and limits red and processed meat, added sugar, refined grains, and starchy vegetables. It provides a framework for dietary transformation that connects chronic disease prevention with sustainable food systems and broader global development goals ^10^. However, translating this framework into cardiovascular prevention requires randomized evidence of its effects on established cardiometabolic risk factors and the biological processes underlying these effects. Comparing the PHD with an ovo-lacto-vegetarian diet also offers an opportunity to evaluate two approaches to predominantly plant-based eating that differ in their inclusion of animal-source foods.

Several PHD indices have been developed to quantify alignment with the EAT-Lancet reference diet, although differences in their construction can limit comparability across studies ^12^. Assessing adherence is essential to interpreting the effects of dietary interventions. However, accurately assessing diet in free-living individuals is complex because of its intrinsic variability over time. Food frequency questionnaires (FFQ) and dietary recalls provide important information on habitual intake but are subject to reporting error and may incompletely capture changes in diet over time ^13^ . Metabolomics offers a complementary approach by identifying circulating metabolites and multi-metabolite signatures associated objectively with dietary intake ^14,15^. Such signatures may strengthen adherence assessment, but their interpretation requires careful distinguishing of biomarkers of food intake from downstream metabolic responses. Although validated biomarkers exist for several key foods and food components relevant to common diets patterns (such as meat, fish, whole grains, legumes), several other important food groups require further biomarker development ^16^. Dietary patterns have also been previously captured through metabolomics, such as the Mediterranean Diet ^17^ and the New Nordic Diet ^18,19^. Extending these efforts to PHD would be valuable to be able to assess adherence in future studies. Alongside accurate assessment of dietary adherence, practical support is needed to help participants adopt and sustain dietary changes. Digital tools, including websites and mobile applications, may facilitate this process by translating dietary recommendations into actionable guidance and providing accesible continous support in daily life, particularly when participants are changing behaviour towards unfamiliar foods and meal patterns ^20^.

The cardiovascular effects of dietary change involve multiple interconnected processes, including lipid metabolism, blood pressure regulation, glucose homeostasis, and inflammation. Responses may vary between individuals due to differences in adherence and underlying metabolic characteristics. Integrating molecular profiling with conventional risk-factor assessment can help characterize these responses and identify biological pathways differentially affected by dietary interventions ^21^. Metabolomics and lipidomics capture changes in circulating small molecules and lipid species, providing insight into metabolic responses to dietary intake ^22,23^. Proteomics provides complementary information on proteins involved in metabolic regulation, inflammatory signalling, and vascular function ^24–26^ . Within a randomized trial, these approaches can identify intervention-responsive molecular features and generate hypotheses about pathways linking dietary change to cardiometabolic health. They may also help characterize heterogeneity in response, informing future precision nutrition research.

Despite growing interest in the PHD and populations becoming more conscious of the need to adopt sustainable diets, important questions remain about its effects on cardiometabolic health. Dietary intervention trials combining conventional cardiometabolic outcomes with emerging omics (i.e., metabolomic, lipidomic, and proteomic profiling) can help determine whether changes in established risk factors are accompanied by broader molecular responses and whether these responses differ between dietary patterns.

The PlanetDiet study is a six-month randomized controlled trial evaluating the effects of sustainable diets on traditional and novel cardiometabolic risk factors in adults at risk of CVD. The primary objective is to test the effects of a sustainable diet on traditional cardiometabolic risk factors, specifically, a metabolic risk score. Secondary objectives are to test the effects of sustainable diets on (1) blood lipids, inflammatory and glycaemic markers, anthropometric measures, and body composition; (2) circulating metabolomic and lipidomic profiles; and (3) circulating proteomic profiles. This protocol describes the study design and methods for evaluating the cardiometabolic and molecular responses to these two dietary interventions. Figure 1 shows the PlanetDiet logo.

**Figure 1.**
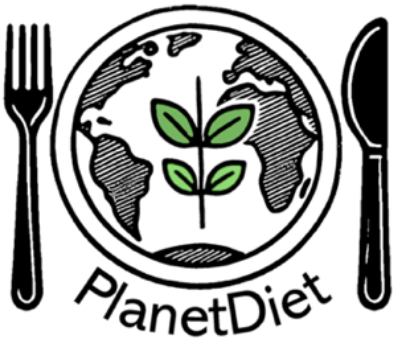
*PlanetDiet* logo. Created with AI (ChatGPT) and Microsoft PowerPoint.

## Methods Study design

The PlanetDiet study is a six-month three-arm parallel RCT in adults (45-70 years old) at CVD risk (n=180, 1:1:1 allocation ratio) (Figure 2). Table 1 illustrates the trial design (PICOTSS strategy). The rationale for selecting participants at cardiovascular risk is that this population is more likely to benefit from preventive dietary interventions, making the findings potentially clinically relevant. Furthermore, these participants may be more prone to changing their diets, facilitating recruitment. A six-month intervention period has been chosen to provide sufficient time for the development of meaningful physiological adaptations ^27^, including changes in cardiometabolic biomarkers, body composition, and dietary habits, while ensuring the feasibility of participant adherence and study implementation. A parallel design has been chosen to prevent potential carryover effects.

**Figure 2.**
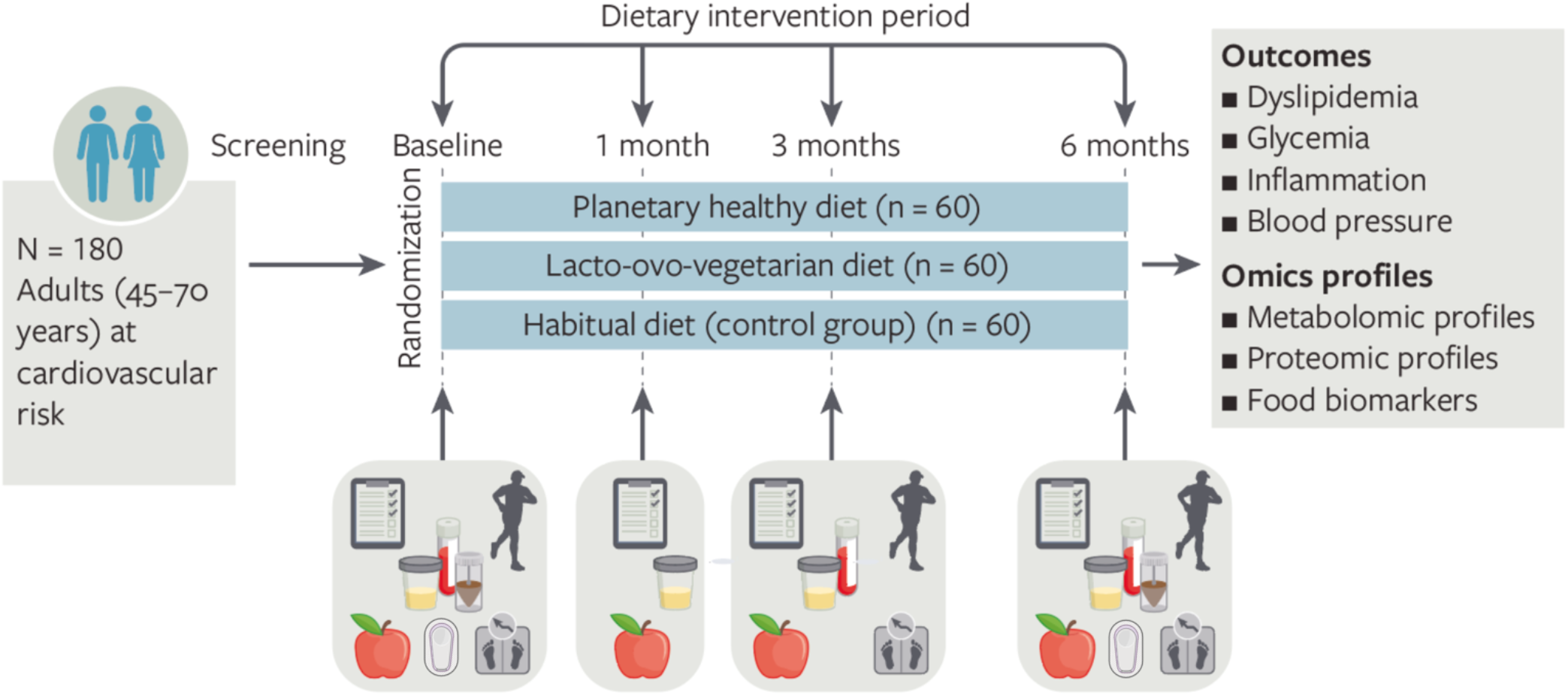
PlanetDiet-overview of investigative approach

**Table 1.** The trial at a glance (PICOTSS strategy)

| **Population** | Men and women (45-70 years old) at cardiovascular risk |
|---|---|
| **Intervention** | Planetary Health Diet and ovo-lacto-vegetarian diet |
| **Comparison** | Habitual diet (general recommendations for adults) |
| **Outcomes** | **Primary outcome:** changes in metabolic health score (triglycerides, HDL-cholesterol concentration, fasting glucose, blood pressure, hs-CRP)<br>**Secondary outcomes**: changes in individual clinical biomarkers (dyslipidaemia, glycemia, inflammation, blood pressure), body composition, metabolomic, lipidomic, and proteomic profiles |
| **Timing** | 6 months |
| **Setting** | Denmark, in a controlled dietary intervention study |
| **Study design** | Randomized controlled parallel-group trial (RCT) |

HDL-cholesterol: High lipoprotein cholesterol, hs-CRP: high-sensitive C-reactive protein.

PlanetDiet study is conducted in accordance with the principles of the Declaration of Helsinki. The protocol, the informed consent, the participant information sheet and the recruitment material have undergone ethical review and approval by the *“The Ethical Committee of the Capital Region of Denmark”*, study code: H-25030722 (date of approval: 18/08/2025). The protocol was prospectively registered at the US National Institutes of Health (ClinicalTrials.gov), registration number: NCT07189676.

## Eligibility criteria for participants

The inclusion and exclusion criteria are described in Table 2.

**Table 2.** Inclusion and exclusion criteria of participants

|  | **Item** |
|---|---|
| Inclusion criteria | Adults (males and females) between 45 and 70 years of age at the time of inclusion. |
|  | Participants must have at least two metabolic alterations: 1) Waist Circumference (WC) >102 cm (males) or >88 cm (females); 2) self-reported medication for blood pressure or blood pressure >130/85 mmHg; 3) self-reported prediabetes or non-fasting plasma glucose 140-199 mg/dL (prediabetes); 4) self-reported lipid-lowering medication or diagnosis of impaired blood lipids (triglycerides: ≥ 150 mg/dL; and HDL: men: < 40 mg/dL and women: < 50 mg/dL). |
|  | Participants are not institutionalized, able to read and provide consent before participation, and willing to attend in-person visits at the study site. |
|  | Participants should have access to a smartphone and computer, or tablet and must be internet-literate. |
|  | Understand Danish both in writing and when spoken. |
| Exclusion criteria | Participants with any serious illness or history of cancer within the past 5 years (except adequately treated localized basal cell skin cancer or in situ uterine cervical cancer). |
|  | Diagnosed with diabetes mellitus, CVD event (myocardial infarction, revascularization procedure, or stroke), or atrial fibrillation. |
|  | Participants with diagnosed psychiatric conditions or cognitive impairment. |
|  | Current smokers including all kinds of nicotine-containing products. |
|  | Body mass index >35 kg/m<sup>2</sup>. |
|  | Known or suspected abuse of alcohol or recreational drugs. Regular alcohol consumption exceeding the Danish national guidelines (i.e., more than 10 standard drinks per week or more than 4 drinks on any single day) will be excluded. |
|  | Pregnancy or planning a pregnancy in the next year. |
|  | Not willing to consume chicken and fish or not willing to make dietary changes related to the intervention. |
|  | Participants with multiple food allergies that could hinder adherence to the intervention. |
|  | Any other issue that makes the project responsible (Principal investigator (PI) or medical responsible) doubt the eligibility of the volunteer. |

### Eligibility criteria for sites and those delivering interventions

Information meetings, individual and group visits take place at the Department of Nutrition, Exercise and Sports (NEXS), University of Copenhagen, Denmark. All study staff involved are trained in handling the equipment and performing the measurements. Dietary intervention is delivered by certified dietitians. The collected biological samples (blood, urine, and faeces) will be analysed after the last participant finishes the intervention by blinded trained laboratory personnel to ensure the consistency of the results.

## Intervention and comparator

### Intervention and comparator description

Participants under free-living conditions are randomized to receive advice on modifying their overall dietary habits according to either a PHD or an ovo-lacto-vegetarian dietary pattern, or to maintain their habitual diet (control group). Participants in the PHD group follow the EAT-Lancet-based diet ^10^ adapted to be nutritionally adequate and culturally acceptable in Denmark, consisting of a high-quality plant-based diet with a limited intake of eggs, dairy, chicken, and fish, but strictly avoiding red and processed meat, processed food and sugars. Participants in the ovo-lacto-vegetarian diet group are advised to adhere to a stricter plant-based diet with intake of eggs and dairy allowed but avoiding chicken, fish, and red and processed meat, and no recommendations on food quantity or the environment foodprint. This comparison will address whether a climate-friendly healthy diet including moderate amounts of animal-based foods provides greater benefits than a stricter ovo-lacto-vegetarian diet ^3^. The two interventions will be compared with the control group, where participants follow their habitual diet without specific recommendations on the consumption of animal products, capturing the general Danish dietary pattern (Table 3). The higher between-group contrast is expected to be found for PHD versus control group.

**Table 3.**
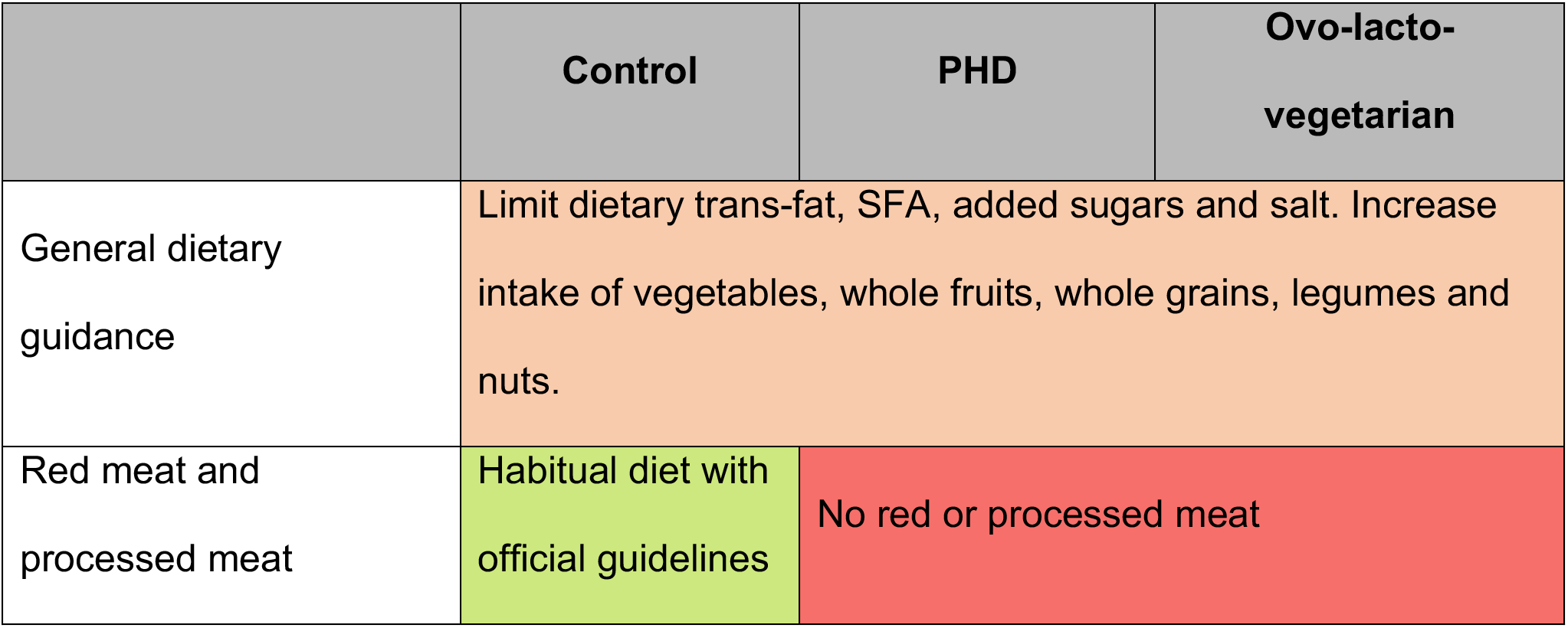
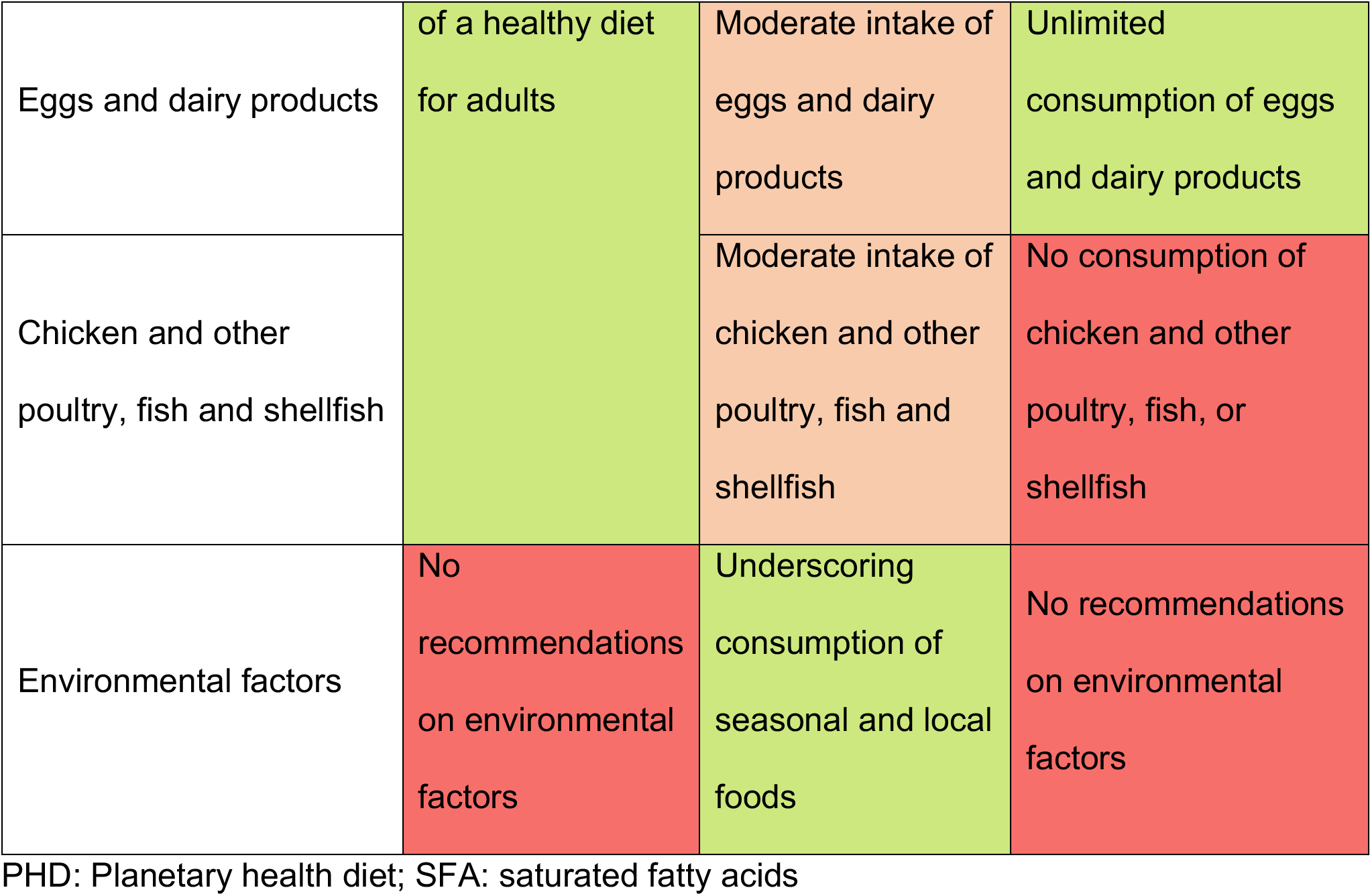
Outline of dietary and environmental guidelines for the three study groups

|  | Control | PHD | Ovo-lacto-vegetarian |
|---|---|---|---|
| General dietary guidance | Limit dietary trans-fat, SFA, added sugars and salt. Increase intake of vegetables, whole fruits, whole grains, legumes and nuts. |  |  |
| Red meat and processed meat | Habitual diet with official guidelines of a healthy diet for adults | No red or processed meat |  |
| Eggs and dairy products |  | Moderate intake of eggs and dairy products | Unlimited consumption of eggs and dairy products |
| Chicken and other poultry, fish and shellfish |  | Moderate intake of chicken and other poultry, fish and shellfish | No consumption of chicken and other poultry, fish, or shellfish |
| Environmental factors | No recommendations on environmental factors | Underscoring consumption of seasonal and local foods | No recommendations on environmental factors |

PHD: Planetary health diet; SFA: saturated fatty acids

Participants allocated to the intervention groups receive detailed dietary guidelines, menus tailored to their assigned diet, guidance on meal planning, and suggestions for substituting meat when preparing meals at home or eating out. In addition, participants receive personalized nutritional counselling during clinical visits with the study dietitians (Supplemental methods 1-4). To support adherence to the intervention, food boxes containing targeted food groups are provided three times during the study period. A specific description and photos of the food content for each group and season can be found in the Supplemental methods 5. Participants are responsible for preparing their own meals at home. Participants are attending two group sessions during the intervention period, providing opportunities to ask questions, receive feedback from study staff, and share experiences with other participants in small groups (5–15 individuals). Specific topics of the group visits as well as all the differences between study intervention groups can be found in Table 4. During these group visits, foods and recipes specific to each study are provided to encourage participants to try new foods and cooking methods.

**Table 4.** Differences in dietary intervention between study arms

| Study arm | Dietary advice | Behavioural support | Food boxes | Group visits |
|---|---|---|---|---|
| **Planetary health diet (PHD)** | 1) Information about the **PHD guidelines (Supplemental methods 1)**; 2) a **meal planner** for their calorie demands; 3) guidance on **legumes intake (Supplemental methods 4).** | 1) **Four individual visits with a dietitian** and; 2) access to a digital **recipe** collection in the study app. Recipes are tagged by **meal type and by season** | **Seasonal** and local fruits and vegetables, **whole grain** products, **legumes, nuts** and **milk alternatives (Supplemental methods 5).** | Topics such as **PHD principles** (**reinforcement of legume intake, reduction of meat intake, guidance on dairy and whole-grain intake), seasonal foods,** and the related **environmental and health outcomes.** Training on how to interpret DXA and CGM results. |
| **Ovo-lacto-vegetarian diet** | 1) Information to adhere to an **ovo-lacto-vegetarian guidelines and** suggestions on alternative for **substituting meat products (Supplemental methods 2)**; 2) guidance on **legumes intake (Supplemental methods 4).** Recommendations on food quantity or on consuming local and seasonal food are not given. | 1) **Four individual visits with a dietitian** and; 2) access to a digital **recipe** collection in the study app. Recipes are tagged by **meal type.** | Fruits and vegetables, **whole grain** products, **legumes, nuts, soy-products, eggs and cheese (Supplemental methods 5)** | Topics such as **plant-based eating and animal-protein alternatives, reinforcement of legumes intake, cooking techniques and nutritional adequacy (iron and protein).** Training on how to interpret DXA and CGM results. |
| **Control (habitual diet)** | Habitual diet + official guidelines of a healthy diet, **without any specific recommendation on animal products (Supplemental methods 3).** | **Four short individual visits with a dietitian**, where they review the recorded dietary data. | Fruits and vegetables, potato, nuts, cheese, **animal protein** products and canned food **(Supplemental methods 5).** | Visits will cover topics such as **the role and importance of controls in RCTs, data protection in clinical trials,** and training on how to interpret DXA and CGM results. |

CGM: Continuous glucose monitor; DXA: Dual-energy X-ray Absorptiometry.

### Criteria for discontinuing intervention/comparator

Study personnel make every effort for the subject to complete the end-of-study assessments as well as document the reason for discontinuation. The Principal investigator (PI) has the right to terminate the participation of any subject at any time if they deem it in the subject’s best interest. Participants lost-to-follow-up and subjects that withdraw either based on their own decision or the investigators will not be replaced. Reasons for early termination include:

1. Personal reasons (e.g. unemployment, moving).
2. Medical reasons (the subject’s condition could contraindicate the study).
3. Non-compliance with the study protocol.
4. Serious Adverse Event (SAE).
5. Lost to follow-up.
6. Other reasons as determined by the investigator.

### Strategies to improve adherence to intervention/comparator, retention and complete follow-up

A study mobile application specifically developed for this study supports participant engagement by providing the study timeline, access to recipes tailored to each study group, instructions for pre-visits tasks such as urine and faecal sample collection, access to self-reported questionnaires to be completed through the app and direct linkage with the dietary record platform for recording 4-day dietary intake. The application also counts with a self-booking system that allows participants to schedule their study visits, with automated appointment reminders. During the trial, participants receive four emails informing them that they can book the next visits. A task reminding them to book attendance will also appear on the app dashboard. The application, named *PlanetDiet and* developed by Unikk.me ApS, is compatible with iOS and Android and is installed on participants’ smartphones during the screening visit (Figure 3).

**Figure 3:**
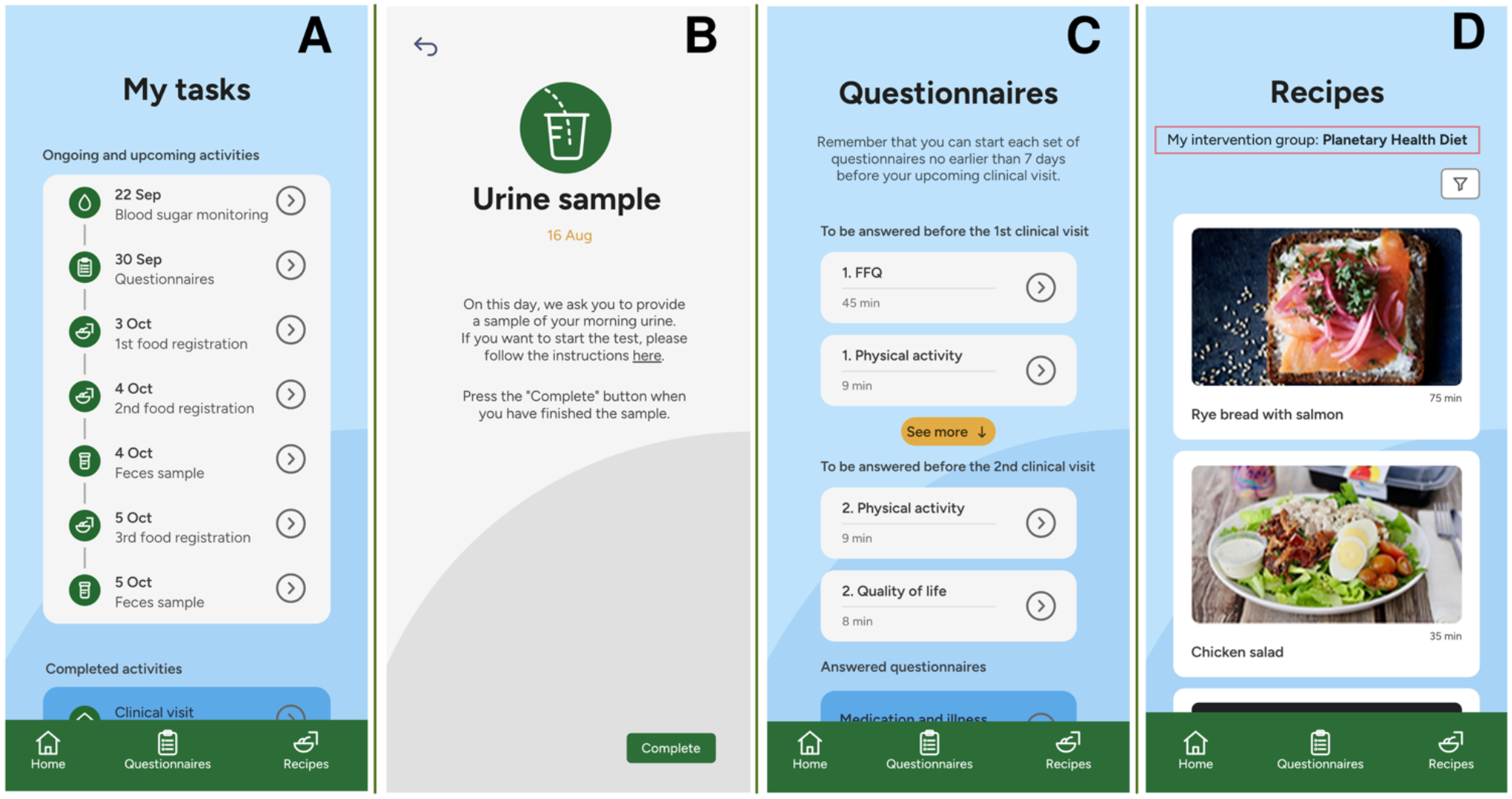
PlanetDiet app. A: task list; B: description of the task; C: self-reported questionnaires; D: recipes tab. App has been designed by Unikk.me ApS (https://www.unikk.me/en). Translated from Danish to English using ChatGPT.

Dietary assessments and intervention’s acceptability data collected through the mobile application are reviewed by study dietitians, who provide individualized feedback to support adherence. Group visits support dietary behaviour change by combining expert guidance from dietitians and researchers with peer-to-peer learning among participants.

Participation is voluntary and without monetary compensation. Participants receive breakfast during clinical visits, home-delivered food boxes (up to 400 DKK (53.5 Eur) per box; Mydietpal ApS), dietitian consultations, and results from clinical assessments, including anthropometry, dual-energy X-ray Absorptiometry (DXA)-based body composition, and 14-day CGM profiles. At the end of the study period, all study participants will be offered the dietary intervention materials for both the PHD and the ovo-lacto-vegetarian diet. Deviations from the standard procedures will be documented.

### Concomitant care permitted or prohibited during the trial

Participants are instructed not to make any lifestyle changes during the six-month intervention, which includes changes in physical activity or over-the-counter medication (unless instructed by the doctor). All concomitant medications that are necessary for the participants’ health and that are not in the protocol exclusion criteria will be continued during the study. The present RCT is not a weight loss trial, thus no total calorie restriction will be advised, and physical activity will not be promoted. A maximum alcohol consumption of 10 drinks/week and no more than 4 drinks on an any single day for men and women drinkers will be allowed.

## Outcomes

### Primary Outcome

The primary endpoint of the trial is the change in a composite metabolic health score from baseline to 6 months. This continuous score is constructed using key clinical biomarkers from main pathways associated with cardiovascular disease, assessed at baseline (V1), 3 months (V3), and 6 months (V4). The specific measurement variables integrated into the score are triglycerides, high-density lipoprotein cholesterol (HDL-c), fasting plasma glucose, systolic blood pressure, diastolic blood pressure, and high-sensitive C-reactive protein (hs-CRP).

The method of aggregation involves calculating an individual Z-score for each continuous variable based on the study population distribution. Standardized values for HDL-c will be multiplied by -1 to ensure all components are directly proportional to cardiometabolic risk. The final composite metabolic health score is determined as the mathematical average of these six standardized component Z-scores, yielding a continuous variable with a mean of 0 and a standard deviation of 1, where higher scores indicate greater cardiometabolic risk. Other researchers have previously applied this approach with some modifications ^28,29^. The primary analysis metric evaluates changes in this continuous score during 6 months of follow-up across the intervention groups. Additionally, to examine non-linear effects, the composite score will be analysed in tertiles.

### Secondary Outcomes

**a) Individual clinical, metabolic, and inflammatory biomarkers:** These endpoints track changes in individual cardiometabolic risk factors using blood and physiological measurements gathered at baseline, 3 months, and 6 months. The specific measurement variables include blood lipids (total cholesterol, low-density lipoprotein cholesterol [LDL-c], HDL-c, triglycerides, and apolipoproteins), glycemia markers (fasting glucose, insulin, and haemoglobin A1c [HbA1c]), and inflammatory markers (hs-CRP, interleukin 6 (IL-6)). The primary analysis metric for these individual markers is the final six-month post-intervention value, adjusted for the baseline value as a covariate using linear regression models. Continuous glucose monitor (CGM) parameters, captured via a 14-day sensor (FreeStyle Libre Pro iQ; Abbott Diabetes Care, Alameda, CA, USA) at baseline and 6 months, will be aggregated into standardized variables including time-in-range and glycaemic variability measures, summarized using mean ± standard deviation (SD) or median and interquartile range (IQR) based on normality.
**b) Blood pressure, anthropometrics and body composition:** Changes in body weight, body mass index (BMI), waist circumference (WC), and systolic/diastolic blood pressure and heart rate are assessed longitudinally at all four clinical visits (baseline, 1 month, 3 months, and 6 months). Body composition variables—including percentage of fat and lean mass—are evaluated via Dual-energy X-ray Absorptiometry (DXA) scans at baseline, 3 months, and 6 months. The analysis metric for these parameters focuses on the longitudinal repeated-measures profile over the six-month period.
**c) Plasma metabolomics and proteomics:** Large-scale molecular profiling are assessed via plasma samples collected at baseline, 3 months and 6 months. Specific variables include targeted and untargeted small-molecule metabolomic and lipidomic profiles using Liquid chromatography–trapped ion mobility spectrometry–time-of-flight mass spectrometry (LC-TIMS-TOF-MS), alongside high-throughput proteomic screening via the Olink Explore HT platform. Approximately 400 metabolites, including amino acids, acylcarnitines, tricarboxylic acid cycle metabolites, bile acids, purines, and pyrimidines, and 300 lipids will be annotated. The Olink Explore HT analysis measures 5416 proteins using 2mL of sample volume with a specificity of 99.5% and negligible cross-reactivity. The analysis metric evaluates six-month post-intervention changes in relative concentrations of metabolites and protein expression levels. Individual proteins will be aggregated into relative concentration units on a log2 scale to generate Normalized Protein eXpression (NPX) values. To handle high dimensionality, metabolites and proteins will also be aggregated into coregulatory modules by applying hierarchical clustering within a Weighted Gene Co-expression Network Analysis (WGCNA) framework to yield module eigenmetabolites and eigenproteins.

### Exploratory Outcomes

Exploratory outcomes capture dietary compliance, behavioural changes, and broad health indicators at baseline, 3 months, and 6 months. The primary analysis metric is the change from baseline to the 3-month and six-month marks. Continuous exploratory metrics will be aggregated and summarized as mean ± SD for normally distributed variables or as median and IQR for skewed distributions and evaluated across intervention groups.

**a) Dietary intake measures:** Dietary variables include 4-day food records (processed via the validated online Myfood24 system ^30^ to track intake of major food groups, energy, saturated fatty acids, total fat, total sugars, fibre, and sodium), and a 200-item FFQ at baseline ^31^. Carbon dioxide equivalents of foods will be obtained using a carbon footprint database based on existing life cycle assessment data. Physiological markers include 3-day faecal samples, bone metabolism markers (C-Terminal Telopeptide of Type I Collagen (CTX), Procollagen Type 1 N-Terminal Propeptide (P1NP)), nutritional status (protein, albumin, ferritin, calcium, vitamin B9 and B12, 25-hydroxy-vitamin D3 concentration), cardiac function (Pro-B-type Natriuretic Peptide (proBNP)), and general liver and kidney function panels. Environmental footprint variables (greenhouse gas emissions, water footprint, CO_2_ equivalents, and land use) will be calculated utilizing life cycle assessment metrics mapped to the dietary intake data.
**b) Biomarkers of food intake:** Biomarkers characterizing specific food groups (e.g. carnosine and anserine for meat, trimethylamide N-oxide, arsenobetaine, and 3-carboxy-4-methyl-5-propyl-2-furanpropanoic acid for fish, 5-dihydroxybenzoic acid and derivatives for wholegrains, hypaphorine for pulses and nuts, daidzein for soy, etc.) will be analysed in spot urine samples, to assess compliance with the targeted food groups ^32,33^ . Moreover, metabolomics signatures of adherences to PHD will also be developed. The urine samples will be analyzed with an LC-Orbitrap-MS Thermo Exploris.
**a) Acceptability and quality of life:** General well-being will be evaluated using the Short Form Health Survey (SF-12) ^34^ which captures both physical and mental health dimensions. Dietary acceptability will be assessed using a Food Acceptability Questionnaire. This self-administered survey evaluates perceptions of feasibility, palatability, and satisfaction with the assigned diet using a 7-points Likert scale ^35^. For the SF-12, we will compute the Physical Component Summary (PCS-12) and Mental Component Summary (MCS-12) using the standard scoring algorithm.

## Participant timeline

The study includes an information visit (1 hour), a screening visit (visit 0, V0) (1 hour), a CGM placement appointment (only when the CGMs cannot be placed during V0 (5 min)), a baseline (visit 1, V1) (2 hours), 1 month (visit 2, V2) (1 hour), 3 months (visit 3, V3) (2 hours), a CGM placement appointment approximately 14 days before the final visit (5 min), and 6 months (visit 4, V4) (2 hours) visit plus two additional group visits (group visit 1, GV1, and group visit 2, GV2) with other participants in the same intervention group (90 minutes). The participant timeline is described in Table 5.

**Table 5.**
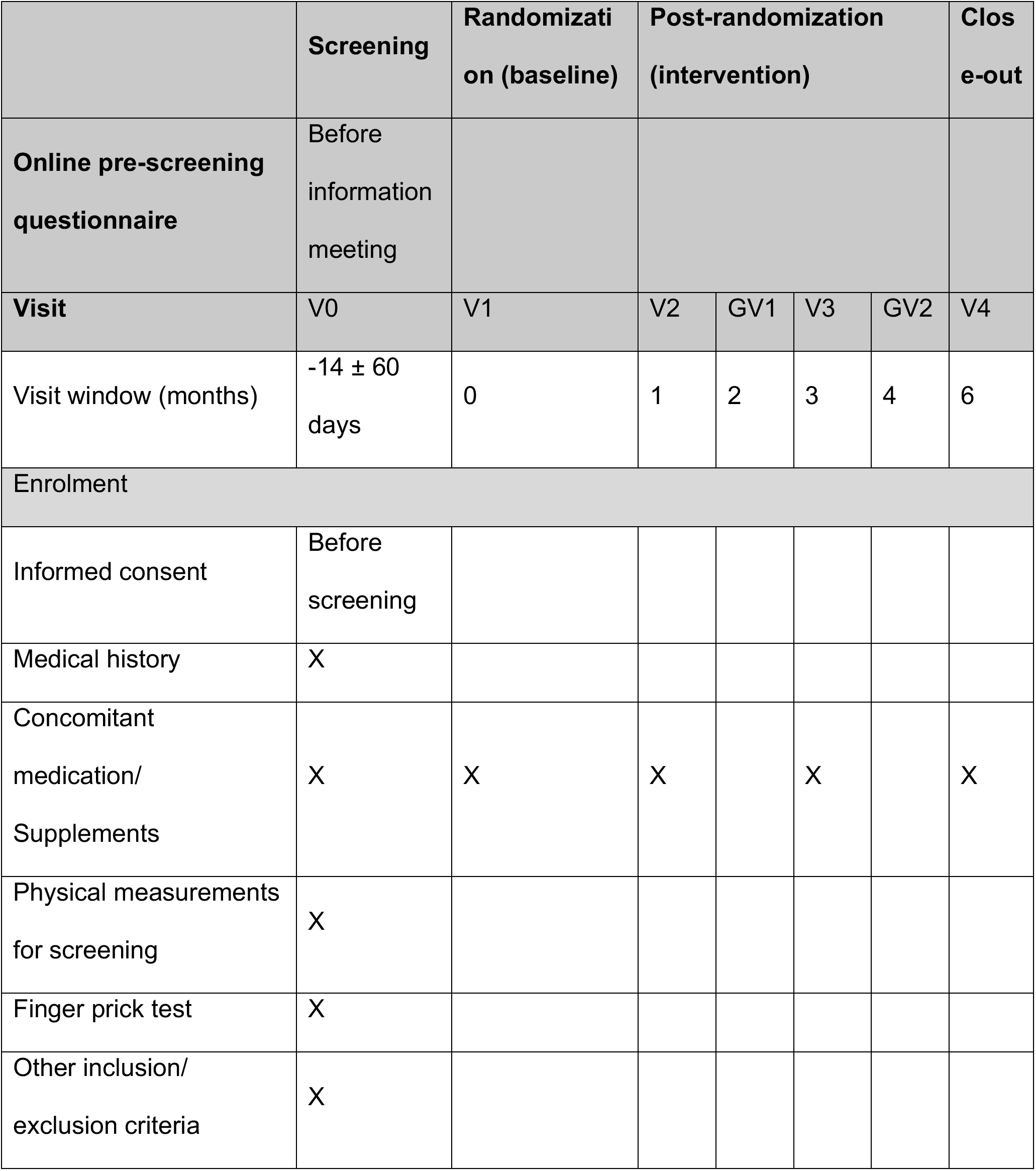
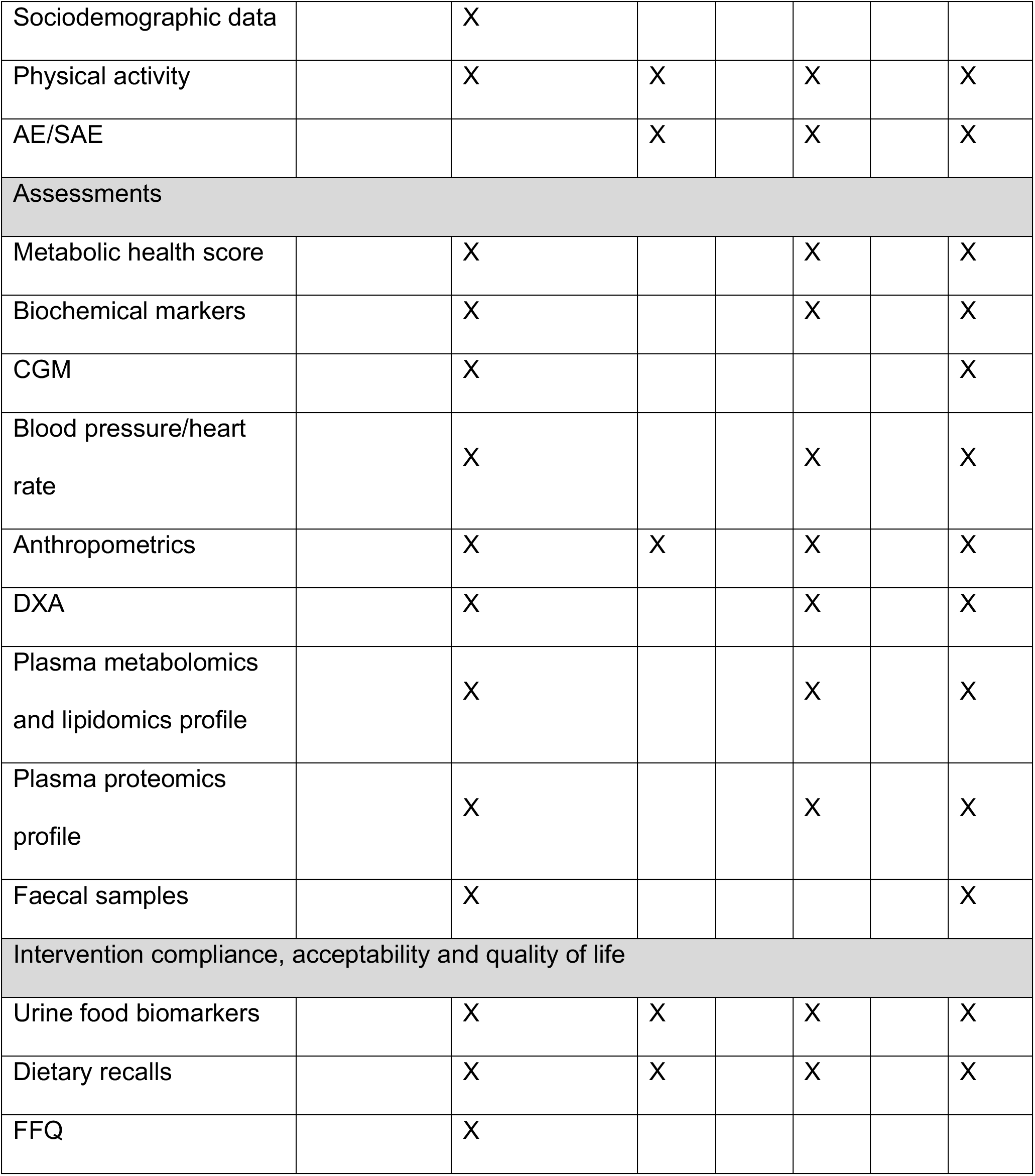
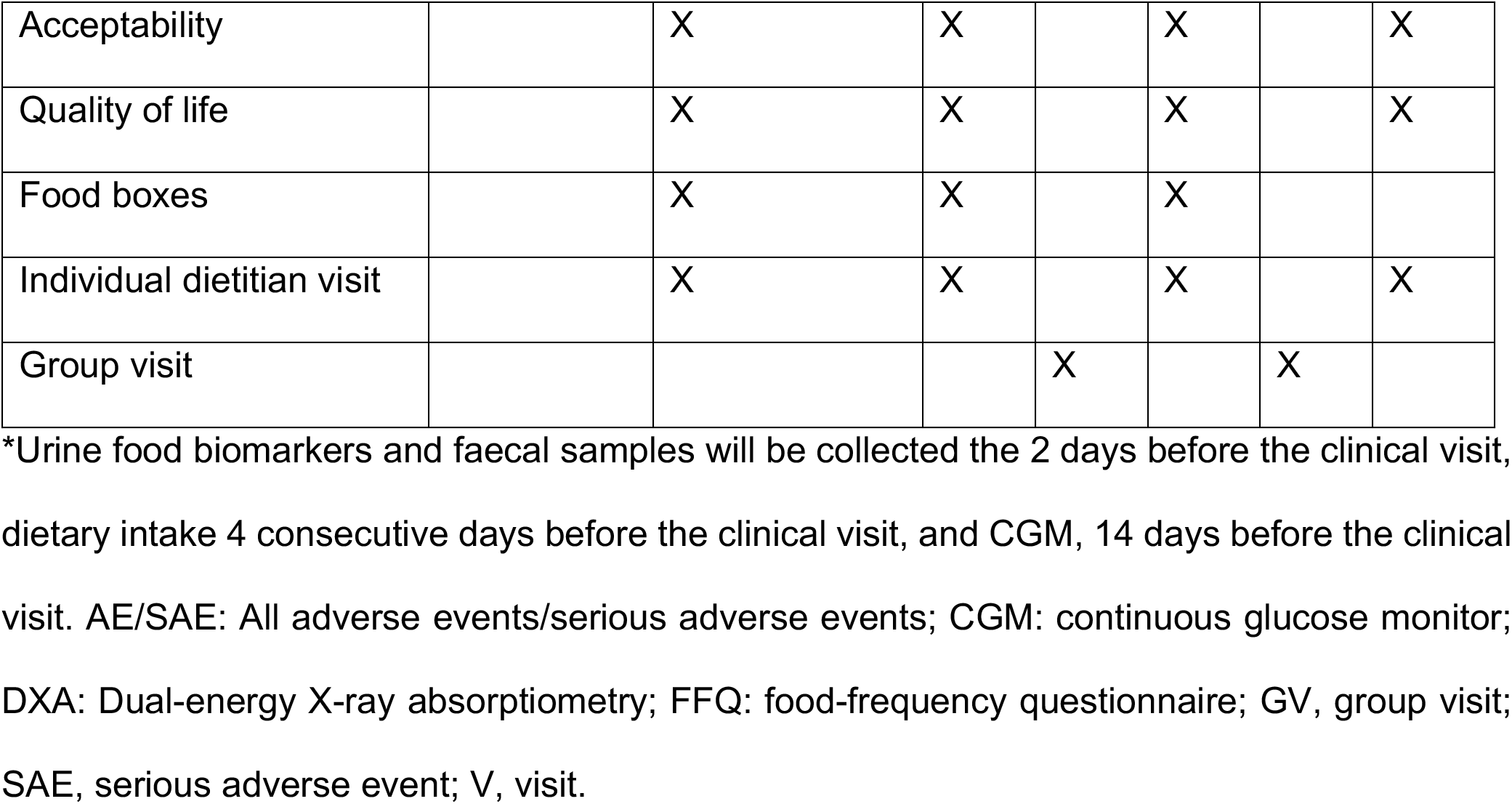
Participant timeline: schedule of enrolment, assessments, and intervention

|  | **Screening** | **Randomization (baseline)** | **Post-randomization (intervention)** |  |  |  | **Close-out** |
|---|---|---|---|---|---|---|---|
| **Online pre-screening questionnaire** | Before information meeting |  |  |  |  |  |  |
| **Visit** | V0 | V1 | V2 | GV1 | V3 | GV2 | V4 |
| Visit window (months) | -14 ± 60 days | 0 | 1 | 2 | 3 | 4 | 6 |
| Enrolment |  |  |  |  |  |  |  |
| Informed consent | Before screening |  |  |  |  |  |  |
| Medical history | X |  |  |  |  |  |  |
| Concomitant medication/ Supplements | X | X | X |  | X |  | X |
| Physical measurements for screening | X |  |  |  |  |  |  |
| Finger prick test | X |  |  |  |  |  |  |
| Other inclusion/ exclusion criteria | X |  |  |  |  |  |  |
| Sociodemographic data |  | X |  |  |  |  |  |
| Physical activity |  | X | X |  | X |  | X |
| AE/SAE |  |  | X |  | X |  | X |
| Assessments |  |  |  |  |  |  |  |
| Metabolic health score |  | X |  |  | X |  | X |
| Biochemical markers |  | X |  |  | X |  | X |
| CGM |  | X |  |  |  |  | X |
| Blood pressure/heart rate |  | X |  |  | X |  | X |
| Anthropometrics |  | X | X |  | X |  | X |
| DXA |  | X |  |  | X |  | X |
| Plasma metabolomics and lipidomics profile |  | X |  |  | X |  | X |
| Plasma proteomics profile |  | X |  |  | X |  | X |
| Faecal samples |  | X |  |  |  |  | X |
| Intervention compliance, acceptability and quality of life |  |  |  |  |  |  |  |
| Urine food biomarkers |  | X | X |  | X |  | X |
| Dietary recalls |  | X | X |  | X |  | X |
| FFQ |  | X |  |  |  |  |  |
| Acceptability |  | X | X |  | X |  | X |
| Quality of life |  | X | X |  | X |  | X |
| Food boxes |  | X | X |  | X |  |  |
| Individual dietitian visit |  | X | X |  | X |  | X |
| Group visit |  |  |  | X |  | X |  |

\*Urine food biomarkers and faecal samples will be collected the 2 days before the clinical visit, dietary intake 4 consecutive days before the clinical visit, and CGM, 14 days before the clinical visit. AE/SAE: All adverse events/serious adverse events; CGM: continuous glucose monitor; DXA: Dual-energy X-ray absorptiometry; FFQ: food-frequency questionnaire; GV, group visit; SAE, serious adverse event; V, visit.

## Adverse effects

At every subsequent study visit, participants are asked if they have experienced any new medical condition since the last evaluation. All adverse events (AE) and serious adverse events (SAE) will be registered in the AE form, including information about the onset of the AE/SAE, end, intensity, relationship to the study product, action taken, outcome and assessment of causality of the AE. Because the trial involves only minor dietary modifications within the framework of healthy dietary patterns and natural foods, the risk of SAE is expected to be very minimal. The investigation team will rate the maximum intensity of each AE reported by a study subject. The intensity grades are defined as follows: a) mild: the AE does not interfere with the subject’s usual function; b) moderate: the AE interferes to some extent with the subject’s usual functions; c) severe: the AE significantly interferes with the subject’s usual function. Volunteers are informed about their right to compensation if they experience minor or major adverse effects because of the study intervention (patienterstatningen.dk). During their participation at NEXS facilities, participants are insured by the University of Copenhagen insurance. Participants will also be covered by the Law on Complaints and Compensation Rights within the Health Care System (Lov om klageog erstatningsadgang inden for sundhedsvæsenet, LBK nr. 962 af 16/08/2024) throughout the study.

## Sample size

To ensure a two-sided paired comparison with 5% significance and 80% statistical power, 60 participants per group will be recruited to detect a difference between groups in the metabolic health z-score. This calculation is based on a previous RCT with similar characteristics that reported a significant reduction of 2.5% in the 10-year Framingham Risk Score between a green-Mediterranean diet and a habitual diet after six months. The common SD was 4.5%, and a dropout rate of 20% has been anticipated in our calculations ^36^.

## Recruitment

Subjects will be recruited from 1) advertisement on social media, University of Copenhagen webpage or similar webpages, as well as printed flyers, 2) voluntary registration via the study webpage, and 3) email invitations sent to individuals listed in the local institutional database who have previously consented to being contacted about future research studies.

On the PlanetDiet webpage, interested individuals can complete a pre-screening indicating their expression of interest, initial consent and a brief inclusion and exclusion questionnaire (Table 6). After this step, potential participants receive the participant information sheet and the official Scientific Ethics Committee’s pamphlet by digital letter. They subsequently can schedule an appointment for an oral information meeting. Potential participants are encouraged to review the materials carefully and may bring a bystander to the meeting. Information meetings take place in a pre-booked interruption-free room and include up to 12 potential participants. After the information meeting, an individual screening visit is performed, including assessment of motivation, medical history, medication use, physical examination, and verification of inclusion and exclusion criteria. The study completion of 180 participants is expected to conclude by October 2027. Recruitment began on 24 November 2025.

**Table 6.** Online pre-screening questionnaire (translated from Danish)

| **Question (answer options)** | **Correct answer for eligibility** |
|---|---|
| Are you between 45-70 years old (Yes/No) | Yes |
| Do you have one or more cardiovascular risk factor (e.g., overweight, high blood pressure, high glucose, high cholesterol) (Yes/No) | Yes |
| Do you smoke or currently use nicotine products (Yes/No) | No |
| Have you been diagnosed with diabetes mellitus, cardiovascular disease or atrial fibrillation (Yes/No) | No |
| Are you willing to follow a diet that includes chicken, fish, or vegetarian diet as part of the trial (Yes/No) | Yes |
| Do you understand Danish both written and spoken (Yes/No) | Yes |

### Informed Consent and Additional Consent Provisions

Potential participants receive two informed consent forms (ICF): (1) An ICF covering participation in the trial and (2) a separate ICF covering General Data Protection Regulation (GDPR) information and the provision of excess biological material to the Copenhagen University’s Biobank for Experimental Research (CUBE) for future research. The ICFs are signed digitally by the participant and the PI or delegated study staff. A copy of the completed ICFs will be sent to the participant via Denmark’s secure digital mailbox. Participation is voluntary, and participants may withdraw their consent at any time without explanation or consequences. Data collected before withdrawal may still be used in the overall analysis.

Participation in the study does not require consent to CUBE biobank storage, and no additional sampling will be collected for this purpose. Participants may also withdraw their consent for CUBE biobank storage at any time before the samples and associated data have been fully anonymized (after 10 years). Any use of the stored samples for future clinical trials will require approval from the relevant Ethics Committee.

## Randomisation and blinding

Randomization was computer-generated by an independent researcher not involved in the trial using Sealed Envelope. Block randomization was used, with blocks of 15 participants, stratified by sex (male/female) and age (45–55/56–70 years). The allocation sequence was uploaded to REDCap. Participants are randomized in a 1:1:1 allocation ratio to one of three groups: PHD, ovo-lacto-vegetarian, or control, through REDCap.

Randomization is performed at the baseline visit to ensure that neither staff nor participants know the assigned group prior to the visit, which could have influenced sampling procedures or measurements. The random allocation sequence is concealed and inaccessible to study staff responsible for enrolling participants and assigning groups. Due to the nature of the intervention, neither the participants nor researchers are blind to the intervention, but laboratory personnel are.

## Data collection and management

### Plans for assessment and collection of outcomes

All clinical visit data are collected using electronic Case Report Forms (eCRFs) in REDCap, hosted by the University of Copenhagen, and entered directly by study staff during visits. Automated range and format restrictions are applied to minimize data entry errors. Self-reported questionnaires, including the 200-item semi-quantitative FFQ ^31^, validated 7-item International Physical Activity Questionnaire – Short Form (IPAQ-SF) ^37^, SF-12 ^34^ and a food acceptability questionnaire ^35^, are completed by participants through the *PlanetDiet* app or participant dashboard webpage (https://planetdiet.dk). Responses are reviewed and, if needed, clarified with participants during study visits. Supplemental Table 1 shows the type of data that is collected, as well as their origin, place of storage, content, estimated size and format.

Clinical markers measured biosamples will be assessed after study completion date by an external laboratory facility blinded to participants’ study allocation and relevant participant data. External partners have access only to the data specified in the data processing agreement. All measurements and sample collections follow established standard operating procedures (SOPs). Data entries and sample handling are regularly monitored to ensure accuracy.

### Data management

The Data Management Plan (DMP) documents the applicable data-management and security procedures and any project-specific adaptations. All data is collected using standardized, predefined questionnaires to ensure consistency across participants. Data is exported to the secure shared storage location every six months. Participants who do not consent to donating excess biological material to CUBE will have all remaining samples destroyed at the end of the study in 2032. The research-biobank is registered at the University of Copenhagen. Personal data is processed in accordance with the General Data Protection Regulation (GDPR; Regulation (EU) 2016/679) and the Danish Data Protection Act.

### Confidentiality

All participant data is handled in a pseudonymized format. Study participants are assigned a unique study number (study ID) as well as a secondary unique identifier. Personally identifiable information (e.g., name, address, CPR number, contact information) is stored separately in an ID-log. This ID-log is maintained under strict confidentiality on the secure shared storage location, with access restricted to a limited number of authorized investigators. Participant identities will remain confidential in all publications. Upon completion of the analysis, participants will be informed of their individual results only if they have provided prior consent.

## Statistical methods

Baseline characteristics for each trial arm will be summarized using descriptive statistics (mean and SD, median and 95% confidence interval, or frequencies and percentages, when applicable). The statistical analysis will be conducted using R and R studio (https://www.r-project.org/).

The primary analysis will estimate the effets of assignment to the PHD and ovo-lacto-vegetarian interventions, each compared with the control group, by evaluating between-group differences in mean change in the composite metabolic health score from baseline to 6 months. A linear mixed-effects model will include intervention group, time (baseline, 3 months, and 6 months), and a group-by-time interaction as fixed effects, with a participant-specific random intercept to account for within-person correlations across repeated measurements. The primary contrasts will assess between-group differences in change from baseline to 6 months; corresponding contrasts at 3 months will assess earlier intervention effects. Analyses will follow the intention-to-treat principle, with participants analyzed according to their randomized allocation, regardless of adherence or intervention completion. A secondary per-protocol analysis will include participants who meet prespecified intervention adherence criteria and have outcome data at 6 months. Before conducting the analyses, assumptions of linear regression, including homoscedasticity and normality of the distribution of residuals, will be explored. In cases where these assumptions are not met, suitable non-parametric analyses will be applied. Clinical biomarkers, anthropometric measurements, dietary intake measures and other exploratory outcomes will be analysed using corresponding mixed-effects models. Binary and ordinal outcomes will be analyzed using appropriate generalized mixed-effects models. A comparative statistical evaluation (repeated measures ANOVA with Tukey’s post-hoc test) will be applied. Additionally, sensitivity analyses will be conducted to assess the robustness of the results to potential confounding arising from differences in baseline characteristics, as well as to different approaches for handling missing outcome data. Missing data will be addressed using baseline observation carried forward (BOCF), whereby the baseline value is used for missing follow-up assessments, and multiple imputation based on group allocation and relevant participant characteristics. Statistical significance will be established at a p-value < 0.05 (two-tailed).

Secondary outcomes include intervention effects on metabolites, lipids, proteins and food biomarkers. To examine the effects of the interventions on individual metabolites, proteins, and food biomarkers, we will use linear regression models Y=β0+β1 X+β2 Y0, in which Y is the metabolite or protein levels at 6 months, Y0 is the metabolite or protein levels at baseline, and X is the intervention arm. Prespecified technical covariates, including analytical batch where applicable, will be included. Features measured at additional time points may be analyzed using mixed-effects models. Quality-control, normalization, transformation, and handling of values below detection limits will be specified for each analytical platform. Multiple testing will be addressed using the Benjamini–Hochberg procedure, controlling the false discovery rate at 5% within prespecified omics analysis families encompassing the two intervention-versus-control contrasts.To reduce dimensionality, we will construct metabolite or protein coregulatory networks by applying hierarchical clustering in the Weighted Gene Co-expression Network Analysis using R package WGCNA ^38^. Finally, we will implement semi-supervised pathway analysis and machine learning methods (such as elastic net regressions) to identify metabolomic and proteomic signatures associated with each intervention. Exploratory mediation analysis will be conducted to examine whether multi-omic profiles mediate the effects of the interventions on cardiometabolic health biomarkers. Data-driven preprocessing, feature selection, and model tuning will be performed within the training folds to minimize information leakage. Primarly analysis will be conducted on an intention-to-treat basis. Secondary analysis will be run stratified by sex.

Univariate (logistic regression) and multivariate (partial-least square discriminant analyses) statistics will be used to assess compliance with the study arms. Logistic regression will be used for binary outcomes (qualitative intake, yes/no) to the intake of targeted food groups and random subject effects will be included to account for individual variability. In parallel, validated predictive models will discriminate meat intake from vegetarians to monitor compliance to the vegetarian diet specifically ^32^. Further, partial least square discriminate analysis (PLS-DA) models will be fitted on the whole panel of food biomarkers comprising the key foods of the three diets to identify non-compliant participants. To develop key signatures for the intervention diets, PLS-DA will be employed, with cross-validation, to select variables based on their importance of projection and selectivity ratio ^18^. Potential determinants of food biomarkers will be analyzed with a linear mixed model including physiologic predictors as fixed effects and subject-specific random effects. Spearman’s rank correlations and partial correlations will be calculated to examine associations between biomarkers, dietary intake, and clinical measures, adjusting for covariates. The key biomarker-based signatures will be validated in external observational cohorts.

For the SF-12, we will compute the PCS-12 and the MCS-12 using the standard scoring algorithm. Mixed-effect linear models with fixed effects for group, time and group X time interactions, and a random intercept for participant to account for repeated measurements. Time will be treated as a categorical variable with four levels: baseline, 1 month, 3 months, and 6 months. For questionnaire items related to acceptability (7-point Likert scale), responses over time across the three intervention groups will be analyzed using cumulative link mixed models, with fixed effects for group, time, and group X time interaction, and a random intercept for participants to account for repeated measures.

For the preliminary descriptive analysis presented in this protocol, we used RStudio (version 4.5.2). Baseline data from 96 participants (recruited up to October 1^st^, 2026) has been used. Categorical variables are described by frequency (%), and continuous variables by mean (SD).

## Frequency and plans for auditing trial conduct

As the trial is not of a medical nature, the regulations of *Good Clinical Practice* do not apply. However, audits and inspections by the relevant authorities, including the Institutional Review Board or Independent Ethics Committee, must be permitted. During these audits, inspections and controls, authorized persons will have access to study data and will be subject to full confidentiality. Participants will be informed that these authorities must have access to their personal data.

## Protocol amendments

Any important protocol modifications will be submitted to the relevant Research Ethics Committee (VEK) for approval. The protocol will not be amended until the proposed changes have received the necessary ethical and regulatory approvals. Clinicaltrial.gov will be consequently updated.

## Dissemination policy

All studies and sub-studies will be published in an open access format. No personal data will be shared openly. This includes addresses, mail, phone number, names and CPR. Data will be converted in findable and reusable format.

## Results

To date, 96 participants (80.2% female) have been randomized (PHD=32, V=32, C=32), with a mean age of 60.0 ± 6.6 years, BMI of 29.0 ± 3.3 kg/m², WC of 100.9 ± 9.6 cm, systolic blood pressure of 138 ± 19 mmHg and diastolic blood pressure of 92 ± 10 mmHg. At screening, 94.8% self-reported medication for blood pressure or presented blood pressure >130/85 mmHg; 87.5% presented WC >102 cm (males) or >88 cm (females); 47.9% self-reported lipid-lowering medication or has diagnosis of impaired blood lipids (triglycerides: ≥ 150 mg/dL; and HDL: men: < 40 mg/dL and women: < 50 mg/dL) and 9.4% self-reported prediabetes or non-fasting plasma glucose 140-199 mg/dL (prediabetes) (Table 7).

**Table 7.** Baseline characteristics in a subsample of the PlanetDiet study

| Demographic and clinical characteristics |  |
|---|---|
| Age, years | 60.0 ± 6.6 |
| Females, % | 80.2 |
| BMI, kg/m<sup>2</sup> | 29.0 ± 3.3 |
| WC, cm | 100.9 ±9.6 |
| SBP, mmHg | 138 ± 19 |
| DBP, mmHg | 92 ± 10 |
| Inclusion criteria prevalence, at screening |  |
| Hypertension, % | 94.8 |
| High abdominal fat, % | 87.5 |
| Dyslipidaemia, % | 47.9 |
| Prediabetes, % | 9.4 |

BMI: body mass index; DBP: diastolic blood pressure; SBP: systolic blood pressure; WC: waist circumference.
Categorical variables are described by frequency (%), and continuous variables by mean (SD).

## Discussion

PlanetDiet is a three-arm, parallel-group, six-month RCT evaluating the effects of sustainable dietary patterns on cardiometabolic health by comparing the PHD with an ovo-lacto-vegetarian diet and a habitual-diet control in adults at increased CVD risk. By combining traditional cardiometabolic risk-factor assessment with metabolomic, lipidomic, and proteomic profiling, the study will characterize both cardiometabolic changes and molecular responses to these dietary interventions. The primary composite metabolic health score integrates four major cardiometabolic pathways: lipid profile, blood pressure, glucose homeostasis, and inflammation, to assess intervention effects across multiple dimensions of cardiometabolic health. Assessing two distinct approaches to predominantly plant-based eating within the same population and study setting will provide evidence relevant to their potential role in cardiovascular prevention.

Achieving and maintaining dietary adherence is a major challenge in nutrition trials ^16^. Dietary choices are made multiple times daily and are strongly influenced by social, behavioral, cultural, and environmental factors. PlanetDiet combines regular participant contact, digital support, and food boxes to help participants translate dietary recommendations into everyday practices. More frequent visits during the initial intervention phase allow study staff to identify and address adherence barriers early, while digital tools provide practical guidance between visits. Previous studies have shown that mobile technologies may support adherence in personalized dietary interventions ^39^. Additionally, food boxes complement these measures by facilitating access to foods included in the assigned dietary patterns. Evaluating adherence and acceptability alongside cardiometabolic outcomes will help contextualize the intervention effects and identify challenges relevant to future implementation. Nevertheless, it is important to note that adherence achieved with intensive trial support and tailored dieticians visit may differ from that attainable in routine practice. This is particularly relevant given the limited evidence on personalized, evidence-based dietary advice and subgroup-specific recommendations for individuals at increased CVD risk ^40,41^ .

Accurate assessment of dietary intake presents an additional challenge as dietary assessment in free-living populations is inherently complex due to variability in intake over time. Traditional methods, including FFQ and dietary recalls, are relatively cost-effective and widely used, but are susceptible to measurement error arising from incomplete food lists, inaccurate portion estimation, recall bias, and random reporting errors ^42^. Food-derived biomarkers measured in blood and urine provide a more objective approach and, together with metabolomics, have enabled the identification of dietary signatures for specific foods and food groups ^16^ . However, biomarkers that capture adherence to predominantly plant-based dietary patterns remain limited. The PlanetDiet trial will therefore provide an opportunity to identify metabolomic signatures and food biomarkers associated with the dietary interventions and investigate their potential as objective markers of plant-based-dietary adherence. Distinguishing markers of dietary intake from downstream metabolic responses will be important, and newly identified signatures will require independent validation before application in other populations.

The PlanetDiet trial has several strengths. First, the combination of clinical and mechanistic outcomes provides an integrated approach to investigating the effects of dietary change on cardiometabolic health. Secondly, the intensive early follow-up of the study facilitates participant engagement, allows study staff to identify and address barriers to adherence at an early stage, and supports monitoring of intervention implementation. The well-defined intervention components and clear differentiation between study arms further strengthen the internal validity of the trial. Including both PHD and ovo-lacto-vegetarian arms will allow the effects of two distinct approaches to plant-based eating to be evaluated within the same population and study setting. In addition, the study places particular emphasis on participant adherence and acceptability of the proposed dietary changes, providing information on the feasibility and sustainability of dietary interventions in a Danish middle-aged population. Furthermore, integrating clinical phenotyping with molecular profiling may reveal biological changes that are not fully captured by conventional risk markers and generate hypotheses about pathways underlying the observed effects.

Several limitations should also be considered. Although the study cannot determine effects on long-term cardiovascular event incidence and it is designed to assess intermediate cardiovascular makers and not hard endpoints, its six-month (26-week) duration is longer than that of many comparable dietary trials, allowing assessment of changes in intermediate cardiovascular risk markers and their changes over the intervention period. Participation may also be subject to selection bias, as individuals with greater health awareness or higher educational attainment may be more likely to participate, potentially limiting the generalizability of the findings to younger or more diverse populations. However, adults at increased cardiovascular risk represent a substantial proportion of the European adult population, supporting the relevance of the study population ^43^. Dietary intake is partly assessed using self-reported dietary registries and may therefore be subject to misreporting, including under- or over-reporting of food intake and inaccuracies in portion-size estimation. Although dietary biomarkers provide an objective complement to self-reported dietary data, they also have limitations. The selected food biomarkers primarily provide qualitative or semi-quantitative information on exposure to specific foods or food groups and may not accurately reflect the absolute amount consumed. Biomarker concentrations may also be influenced by individual metabolism, timing of sample collection, and other lifestyle or physiological factors. Consequently, dietary registries and food biomarkers should be considered complementary measures rather than direct quantitative measures of dietary intake. Attrition and incomplete longitudinal data may introduce bias, particularly if loss to follow-up or missing data differ between study arms or are associated with intervention adherence. To mitigate the impact of attrition, the planned sample size includes an additional 20% to account for potential dropouts. Adherence to the dietary interventions and completeness of dietary and CGM data may also vary between participants. Data completeness and adherence will therefore be monitored throughout the intervention by the study staff. Predefined quality-control criteria will be applied to identify insufficient data, and sensitivity analyses will be conducted to assess the robustness of the findings to missing or incomplete longitudinal data.

PlanetDiet will contribute randomized evidence on the cardiometabolic effects of dietary patterns intended to support both human health and environmental sustainability. Integrating conventional risk factors, adherence measures, and molecular profiles may inform subsequent trials and help identify dietary response patterns that warrant further investigation. Translation into routine clinical practice and public health programs will require evaluation of long-term adherence, feasibility, resource requirements, and cost-effectiveness, including the support and training needed for healthcare professionals delivering dietary guidance. The findings will inform future research and implementation strategies for incorporating sustainable dietary patterns into cardiovascular prevention.

## Data Availability

The study protocol and statistical analysis plan will be available from the corresponding author upon reasonable request. Following publication of the main trial results, deidentified participant-level data, the data dictionary, statistical code, and selected study materials will be made available upon reasonable request, subject to applicable ethical approvals, data protection requirements, and approval by the study investigators.

## Declarations Acknowledgements

We are grateful to the study staff (Jane Guldborg Jørgensen; Anna Sønder Mogensen; Laura Møller Helmersen; Marie Duemose Krøll Brunsgaard; Emilia Bjergskov Pedersen; Kasper Birkerod Lillholm), and all participants of the PlanetDiet study for their work and support.

## Authors’ contributions

Conception and design: M.T-S, C.C, I.T, and M.G-F; drafting the manuscript: M.T-S, and M.G-F. Revising it critically for important intellectual content: T.C-S, C.C, A.R, AM. R, and I. T; Funding: I.T and M.G-F; material support: T. C-S and AM R; and final approval of the version to be published: all co-authors.

## Ethics approval and consent to participate

PLANETDIET: Sustainable diets and cardiometabolic health: a multi-omics approach in a Randomized Controlled Trial (RCT). The Ethical Committee of the Capital Region Denmark E-Journal-nr.: H-25030722, amendment number 126414.

## Non-standard Abbreviations and Acronyms

AE: Adverse event
BOCF: Baseline observation carried forward
CGM: Continuous glucose monitoring
CTX: C-terminal telopeptide of type I collagen
CUBE: Copenhagen University’s Biobank for Experimental Research
DMP: Data Management Plan
eCRF: Electronic case report form
GDPR: General Data Protection Regulation
GV1: Group visit 1
GV2: Group visit 2
Hs-CRP: High-sensitivity C-reactive protein
ICF: Informed consent form
IPAQ-SF: International Physical Activity Questionnaire – Short Form
LC-TIMS-TOF-MS: Liquid chromatography–trapped ion mobility spectrometry–time-of-flight mass spectrometry
MCS-12: Mental Component Summary
NNF: Novo Nordisk Foundation
NEXS: Department of Nutrition, Exercise and Sports
P1NP: Procollagen type 1 N-terminal propeptide
PHD: Planetary Health Diet
PICOS: Population, Intervention, Comparator, Outcomes, and Study design PLS-DA Partial least squares discriminant analysis
PCS-12: Physical Component Summary
proBNP: Pro-B-type natriuretic peptide
SAE: Serious adverse event
SF-12: 12-Item Short Form Health Survey
SOP: Standard operating procedure
WGCNA: Weighted gene co-expression network analysis

